# The auditory Mismatch Negativity reflects accelerated aging in adults with Down’s Syndrome

**DOI:** 10.1101/2020.12.21.20243188

**Authors:** Chiara Avancini, Sally Jennings, Srivas Chennu, Valdas Noreika, April Le, Tristan Bekinschtein, Madeleine Walpert, Isabel Clare, Anthony Holland, Shahid Zaman, Howard Ring

## Abstract

Down’s Syndrome (DS) is associated with premature and accelerated ageing and a propensity for early-onset Alzheimer’s disease (AD). The early symptoms of dementia in people with DS may reflect frontal lobe vulnerability to amyloid deposition. The Mismatch Negativity (MMN) is a frontocentral component elicited by auditory violations of expected sensory input and it reflects sensory memory and automatic attention switching. In the typically developing (TD) population, the MMN response has been found to decrease with age. In the cross-sectional phase of this study the MMN was used to investigate the premature neurological ageing hypothesis of DS. In the longitudinal phase, we evaluated the MMN as a potential predictor of cognitive decline. The study found that age predicted MMN amplitude in DS but not in those who are TD, showing that the MMN reflects accelerated ageing in DS. However, a follow-up of 34 adults with DS found that neither amplitude nor latency of the MMN predicted cognitive decline one year later.

## 1. Introduction

People with Down’s Syndrome (DS) experience premature and accelerated ageing. This is evident across several physiological systems: from earlier menopause to premature skin wrinkling and early onset of dementia (Zigman, 2013). Alzheimer’s disease (AD) pathology affects nearly the entirety of the DS population as they age with a prevalence of clinical dementia in over 40% by the time individuals with DS reach their 50s (Holland et al., 1998) and 75% by their 60s (Holland & Ball, 2009). In DS there is amyloid overproduction as a result of the triplication of the amyloid precursor protein gene, which is located on chromosome 21. Similarly to the Typically Developing population (TD), cerebral amyloid binding is increased once symptoms of dementia emerge and when clinically diagnosed dementia is apparent. The pathology progression seen in DS resembles that of sporadic AD (Annus et al., 2016). Neuropathological changes associated with Alzheimer’s disease (AD) begin years before symptoms onset (Sperling et al., 2013; Teipel et al., 2020) and therefore, early detection and the ability to track pathological progression has the potential for maximizing the benefits of pharmacological and clinical interventions, ultimately leading to prevention of dementia, increased life expectancy and improved quality of life. Hence, effort has been put into identifying markers that can reliably measure premature ageing and predict future cognitive decline (Bibina et al., 2017; Counts et al., 2017; de Roeck et al., 2019; Sperling et al., 2013).

Positive amyloid or tau tracer binding in positron emission tomography (PET) imaging, low cerebro-spinal fluid (CSF) concentrations of amyloid, high CSF concentrations of total tau and phospho-tau, and MRI measures of cortical and hippocampal structure and function have been suggested to be accurate biomarkers of AD progression (Annus et al., 2016; Handen et al., 2012). While the utility of these measures in early detection and tracing of AD progression is arguably of fundamental importance, they overlook one key feature - the brain is a dynamic system that processes internal and external stimuli in a matter of milliseconds, and it is this ability that ultimately determines cognitive function and behaviour. In order to fully understand AD pathology, it is therefore crucial to understand how AD functionally affects brain dynamics. This can be achieved with electroencephalography (EEG), which also has the advantage of being an inexpensive, non-invasive and relatively participant-friendly technology, making it suitable for using with the DS population. Sensory memory is a function that has been found to decline with typical ageing. This has been assessed electrophysiologically by testing the deterioration of the auditory Mismatch Negativity response (MMN) (Cheng et al., 2013; Horváth et al., 2007; Kiang et al., 2009; Näätänen et al., 2011; Schiff et al., 2008) with age. In the TD population, the amplitude of the MMN is attenuated and the latency slowed in older compared to younger individuals (Cheng et al., 2013; Horváth et al., 2007; Kiang et al., 2009; Näätänen et al., 2011; Schiff et al., 2008). The MMN is a frontocentrally distributed Event-Related Potential (ERP) component peaking at 150-200ms after stimulus onset. It is elicited by the occurrence of violations of auditory regularities encoded as sensory memory trace in the auditory cortex (Näätänen et al., 2007; Näätänen et al., 2004). The presentation of a stimulus that deviates in one or more features from the regular stimuli creates discordance between the incoming information and the memory trace, eliciting the MMN. This occurs even if the participant is not attending to the stimuli, supporting the hypothesis that the MMN functionally reflects an involuntary attention switch to auditory changes (Näätänen et al., 2007; Paavilainen, 2013). The MMN has also been interpreted within the predictive coding framework (Friston, 2003, 2005). Predictive coding postulates that the brain creates neural predictions about future events by compiling statistical regularities from incoming stimuli. The processing of information leading to the formation of statistical models of the outside world is thought to rely on a hierarchy of brains structures and mechanisms (Huang & Rao, 2011). The cortex generates top-down predictions that are then compared with incoming sensory stimuli at the first level, or to a bottom-up input at any higher level. If there is a discrepancy between the top-down prediction and the bottom-up input, a prediction error occurs. Such prediction error is fed back to the higher levels of the processing hierarchy and it is used to update the internal model of the environment. The MMN response corresponds to the prediction error that is modulated by the interplay between the perceptual response contingent on the deviant stimulus and changes in synaptic sensitivity during perceptual learning (Garrido et al., 2008; Garrido et al., 2009b).

The MMN is generated by a temporofrontal network comprising the bilateral auditory cortex (A), the bilateral supratemporal gyrus (STG) and the right inferior frontal gyrus (rIFG) (Garrido et al., 2008, 2009a, 2009b). These cortical sources interact with each other hierarchically by changes in the connectivity within cortical sources and between each other. Initially, the A adapts to repeated standard stimuli. Then, changes in connectivity between A and the frontotemporal nodes of the network reflect changes in the precision of the prediction error that is fed from lower sensory levels to higher levels of the hierarchy responsible for top-down predictions. The MMN results from the failure of the higher levels of the hierarchy to predict the bottom-up input, which results in changes of coupling between the sensory and the frontotemporal cortices (Garrido et al., 2008, 2009a, 2009b). The role of the frontotemporal network in the generation of the MMN has been supported by studies on clinical populations. Specifically, studies on people with frontotemporal dementia have shown that the disruption of frontotemporal connections impairs the detection of change by the auditory cortex (Hughes & Rowe, 2013) and frontal brain injury patients have reduced MMN responses (Alain et al., 1998; Alho et al., 1994).

In DS the frontal lobe is smaller and less developed than in controls, hence the potential vulnerability of the frontal lobe to amyloid accumulation may explain the early onset of executive symptoms in people with DS in the preclinical stages of AD (Annus et al., 2016; Lautarescu et al., 2017; Neale et al., 2018). On the basis that the MMN is generated by frontal and supratemporal cortices, we hypothesised that changes across age in the MMN may reflect accelerated ageing and be associated with age-related cognitive decline in people with DS. Furthermore, the independence that the MMN has from attention makes it a suitable measure in DS who experience impaired attention (Lott & Dierssen, 2010). To investigate this, participants with DS and age-matched TD controls were presented frequent and deviant tones during EEG recording as well as being administered neuropsychological tests sensitive to age-related cognitive decline. After one year, the same participants with DS were re-administered the neuropsychological test. Specifically, we hypothesized that increasing age predicted smaller MMN amplitudes and longer latencies in both groups with a stronger effect in DS compared to TD participants. Furthermore, we hypothesized that the MMN at the first timepoint would predict cognitive decline in DS as measured by comparison in neuropsychological test scores between the time points one year apart.

## 2. Materials and Methods

The present study was conducted with both a cross-sectional and a longitudinal phase. In the cross-sectional phase both DS and aged-matched Typically Developing (TD) participants took part to the EEG session (described in section 2.4) and were administered neuropsychological and cognitive tests (described in section 2.3), as well as having a hearing check. In the longitudinal phase, only the participants with DS were re-tested on neuropsychological tests to determine whether there had been cognitive decline since time point 1 (described in section 2.4).

### 2.1. Participants in the cross-sectional phase

Thirty-six adults with DS aged 20 years or older were recruited into the study. Participants with DS were predominantly identified through their previous participation in the ‘Defeat Dementia in Down’s Syndrome’ research programme. Participants who were not already known to the research group were made aware of the study through adverts by the Down’s Syndrome Association (DSA). Thirty-nine age- and gender-matched TD control participants were also recruited into the study, identified through the Join Dementia Research (JDR) database. Ethical approval to conduct the study (reference 14/LO/1411) was given by the National Research Ethics Service (NRES). The Committee had the expertise to assess studies that might include individuals who lacked capacity to consent to participation in research.

### 2.2. Participants in the longitudinal phase

Thirty-four adults with DS who completed the initial EEG assessment were re-approached 10-14 months later (mean 12 months) for a follow-up cognitive assessment using the participant-based CAMCOG-DS, and for an informant-based diagnostic interview with the parent or carer using the CAMDEX-DS.

### 2.3. Clinical assessments

#### 2.3.1. Hearing checks

As the EEG paradigms involved the presentation of auditory stimuli hearing loss was screened for with the Siemens HearCheck Navigator, which has been validated as an appropriate tool (Fellizar-Lopez et al., 2011). This HearCheck Navigator sequentially delivers tones at two frequencies (1000Hz, 3000Hz) and a range of decibels (20dB – 75dB) and participants who did not hear tones of 1000Hz and 3000Hz at 55dB were to be excluded from the study, although no-one with this degree of hearing loss was identified.

#### 2.3.2. Intellectual Functioning

The Kaufman Brief Intelligence Test-2. (KBIT-2; Kaufman & Kaufman, 2014) was used to estimate intellectual functioning. The KBIT-2provides verbal (VIQ) and nonverbal IQ (NIQ) scores. Normally, these are to produce a Composite IQ score. Where the VIQ and NIQ discrepancy is too large (see Table B 7, *ibid*.), Verbal IQ on its own is used.

#### 2.3.3. Dementia screening in TD

The Addenbrooke’s Cognitive Examination Revised (ACE-R; Mioshi et al., 2006) was used to screen control participants for dementia. Control participants were excluded at the lower cut-off of 88 or below. No control participants were excluded from this study based on their ACE-R assessment.

#### 2.3.4. Cognitive decline in DS

The Cambridge Examination for Mental Disorders of Older People with Down’s Syndrome and Others with Intellectual Disabilities (CAMDEX-DS; Fonseca et al., 2018; Holland & Ball, 2009; Roth et al., 1986) was developed as a tool to aid the diagnosis of dementia in people with intellectual disability. It includes a cognitive assessment component (CAMCOG-DS) and an informant interview (CAMDEX-DS). The CAMCOG-DS assesses seven functional domains affected by the presence of AD: orientation, language, memory, attention, praxis, abstraction, and perception. The total possible score is 109.

The CAMDEX-DS informant interview (Fonseca et al., 2018) was used to identify functional decline and to structure the diagnosis of dementia in people with DS based on reported change across specific functional domains. A dementia diagnosis from the CAMDEX-DS is a clinical decision based on parent or carer reports of the participant’s best level of cognitive functioning and evidence of functional decline. The interviewee is selected on the basis of having had regular contact with the participant for at least six months prior to the assessment. In this study the diagnosis of dementia in a participant with DS was made by an experienced psychiatrist reviewing, blind to the age, gender and previous diagnostic status of the participant, the data collected using CAMDEX-DS informant interview with the parent or carer.

### 2.4. EEG assessment

#### 2.4.1. Task paradigm

A modified version of the auditory global-local paradigm (Bekinschtein et al., 2009) was used, previously described in Chennu et al. (2013). consisting of the presentation of tones at a volume that the participants indicated as audible and comfortable. Each tone lasted 50ms and was presented in group sequences of five tones with 100ms intervals in between each tone. The five-tones group consisted of either local *standard* sequences in which five tones of identical pitch (AAAAA or BBBBB) were presented, or local *deviant* sequences in which the first four tones were identical and the last tone was of a different pitch (AAAAB or BBBBA). The tones themselves were mixtures of three sinusoids of either type: A (500, 1000, and 2000 Hz), or B (350, 700, and 1400 Hz). The tone sequences were presented either entirely monaurally (AAAAA, BBBBB, AAAAB, BBBBA), to the left or right ear, or predominantly monoaurally with the final tone presented on the opposite ear (AAAA*A*, BBBB*B*, AAAA*B*, BBBB*A*. See Fig. 1).. Tone sequences were presented in experimental blocks, and each block included approximately 160 sequences and was counterbalanced by the dominant tone type (A or B) and the laterality of monoaural tone delivery (left or right). There were two block types. In block type X, local *deviant* sequences were 14.25% of the total sequences, the rest being local standards. In block type Y, local *deviant* sequences were 85.75% of the total sequences.

**Figure 1.**
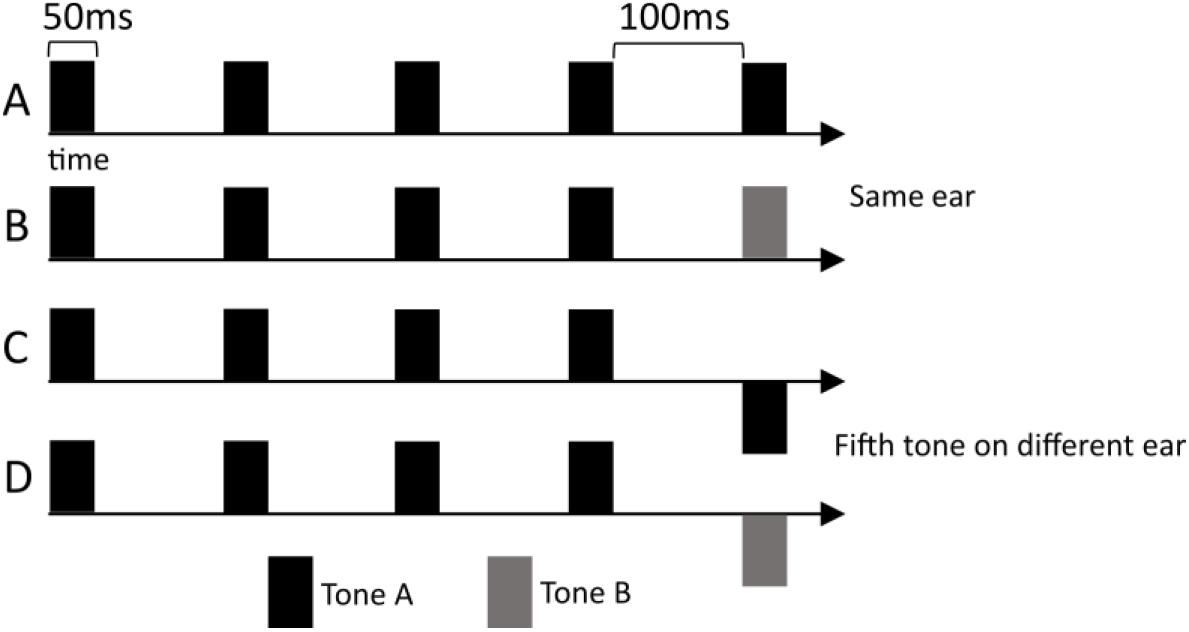
Paradigm: Examples of five tones sequences. In *standard* sequences, tones were all of the same pitch (A and C). In *deviant* sequences, the first four tones were identical and the fifth was of a different pitch (B and D). Sequences could either be presented monoaurally on the same ear (A and B) or interaurally with the fifth tone presented on the opposite ear (C and D). Tones were 50ms long and were spaced by 100ms silence.

At the beginning of the testing session, participants were informed that they were about to hear groups of sounds. Participants were asked to listen carefully to the groups of sounds because at the end of each block they would be asked: “can you tell me what group of sounds you heard a lot?” and “can you tell me what group of sounds you heard sometimes?”. Participants’ answers were recorded at the end of each block. The purpose of the questioning was to maintain participants’ attention on the groups of sounds. At the end of each block, participants were also asked about their arousal levels on a scale of 1-10 (1 – Asleep to 10 – Fully awake), and attentiveness (1 – Mind wandering/unattentive to 10 – Fully attentive to stimuli). Participants took a break between each block at a length of their choosing. A total of 8 experimental blocks (4 Y blocks and 4 X blocks) were presented, with total testing time, including breaks and questioning, taking an average of 40 minutes.

The global-local paradigm is measure prediction error responses to two hierarchical levels of deviation: (i) global – between trial variance: which requires the participants to report rare sequences of tones; (ii) local – within trial variance: the automatic detection of individual deviant tones. As shown in Chennu et al. (2013), the perception of global deviance elicits an attention-dependent P300 response, whereas the detection of local deviance elicits a MMN. This paper is focused on the local effect, which elicits an MMN response, even in the absence of explicit attention.

#### 2.4.2. Data acquisition

Data were recorded with 129-channels EEG gel nets (EGI’s HydroCel Geodesic Sensor Net). Testing was conducted in an electrically shielded room, using the Net Amps 300 amplifier (Electrical Geodesics). The auditory stimuli were presented to participants using Psychtoolbox-3 (Delorme & Makeig, 2004) running on MATLAB; at a comfortable volume as judged by participants; binaurally through Etymotics ER-3A earphones. The EEG data was recorded onto NetStation Version 5(Magstim EGI). The recording parameters for collection were: <100KOhms impedance, 500 Hz sampling rate, and referenced to the vertex.

#### 2.4.3. Data preprocessing

Pre-processing was run in MATLAB using custom functions along with EEGLAB toolbox (Chennu et al., 2013). Channels of the outer circle of the net were excluded from the pre-processing as they carry little neural information and mostly muscle artifacts (Chennu et al., 2013). Continuous data were low-pass filtered offline at 20Hz (Garrido et al., 2008; Jacobsen & Na, 2005; Näätänen et al., 2004). Epochs were then selected from −200ms to 700ms relative to the onset of the fifth tone in each sequence and baseline corrected −200ms to 0ms relative to the 5th tone onset. Bad electrodes were detected by a quasi-automated procedure: noisy channels were identified by calculating their normalized variance and then manually rejected or retained by visual inspection. Rejected channels were excluded from ICA decomposition, which was used to remove eyeblinks and lateral saccades. Robust detrending was applied on the ICA corrected signal to correct for slow drifts. Subsequently, rejected channels were interpolated and epochs exceeding ±150µV were marked and then discarded following further visual inspection and the signal was then re-referenced to the average. An average of 6% of the trials were rejected (range = 0-36%). The Global Field Power (GFP) and the ERP was obtained for *standard* and *deviant* conditions and the MMN *difference waves* (dERP and dGFP) were obtained by subtracting the waveform of *standard* epochs from the waveforms of *deviant* epochs and were baseline corrected after the subtraction. The GFP was obtained by calculating the point-by-point standard deviation of the ERP voltage and was then baseline corrected.

Because the MMN has not yet been characterised in DS, the peak of the MMN was initially identified by the inspection of butterfly plots (Fig. 2) and thereby identified as occurring at 140ms after stimulus onset. In order to select the relevant electrodes for ERP analysis for each group, the cluster-permutation algorithm (run with FieldTrip toolbox; Maris & Oostenveld, 2007) was run separately for controls and DS contrasting *deviant* and *standard* trials. Two-tailed dependent t-tests were used to evaluate the effect. At the cluster level, the null distribution was generated using the Monte Carlo method and the critical value used for thresholding the sample-specific T-statistics was set at α = 0.01. Separately for both groups, the relevant electrodes to be kept for further analysis were selected as the electrodes forming a significant negative cluster between 120ms and 160ms after stimulus onset (Fig. 3) and the ERP MMN peak detection was performed on the average of the selected electrodes. For each individual participant the peak corresponding to the MMN was defined as the most negative value between 120ms and 160ms for the ERP of *standard* and *deviant* trials and the dERP. For the GFP of *standard* and *deviant* trials and the dGFP, the MMN was defined as the most positive value between 120ms and 160ms. Both amplitudes and latencies were extracted for statistical analyses.

**Figure 2.**
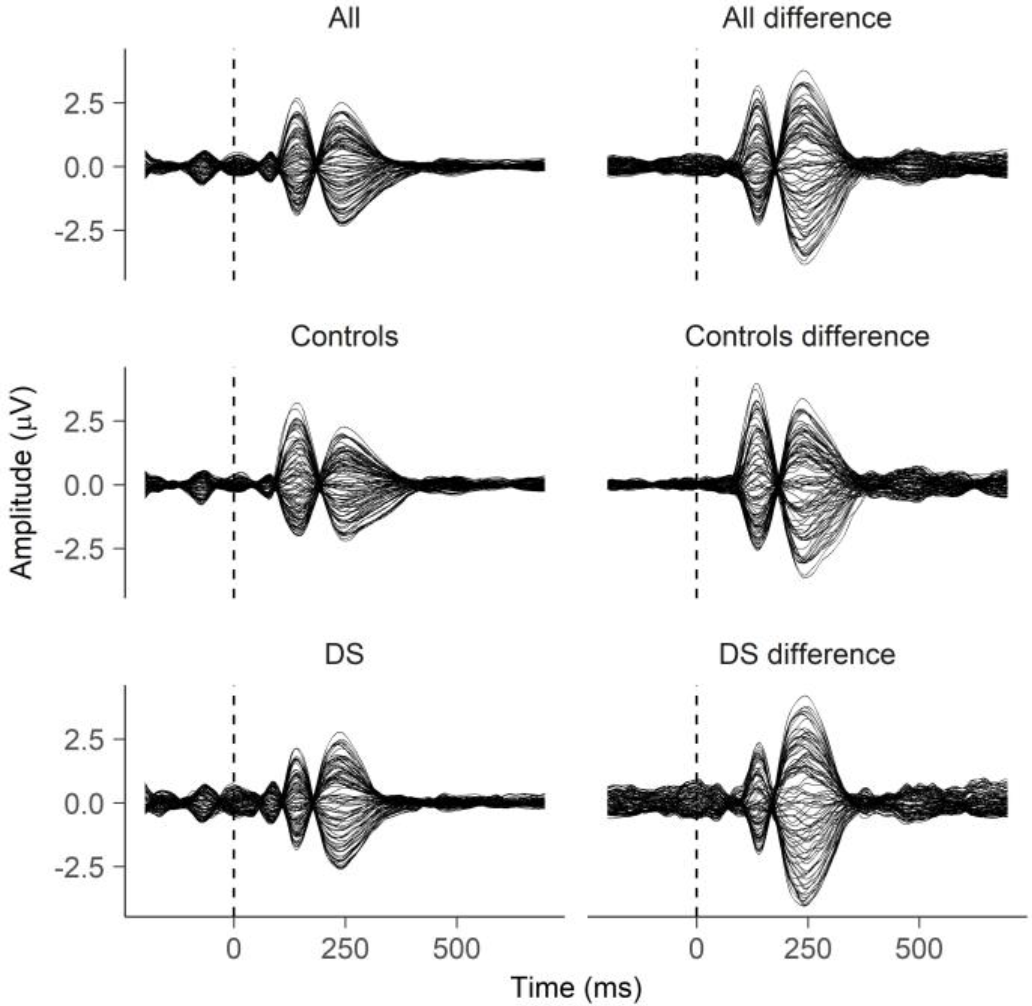
Butterfly plots: Grand average of all electrodes in DS, controls and all participants for both *standard* and *deviant* conditions combined (first column). ERP of all electrodes in DS, controls and all participants of the *difference waveforms* (second column).

**Figure 3.**
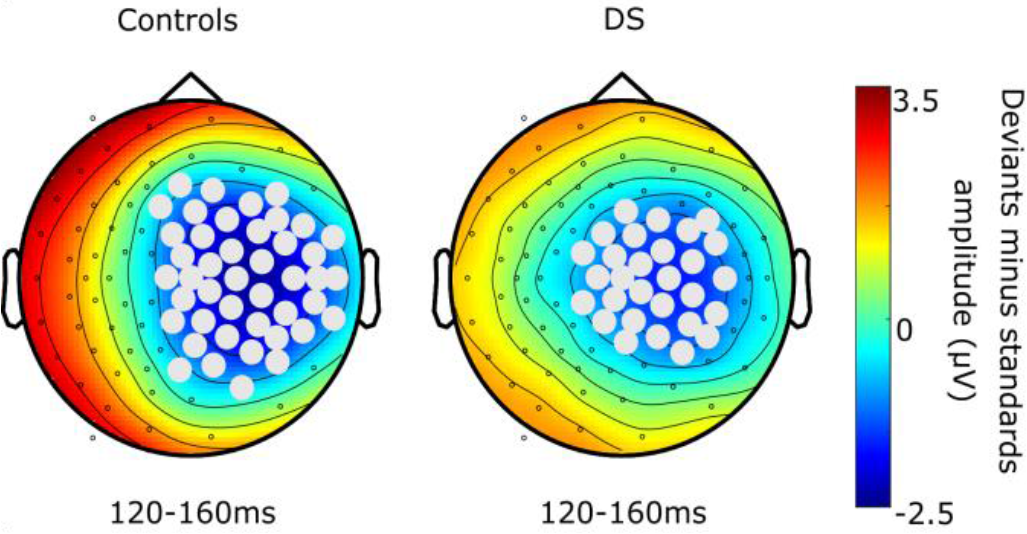
MMN electrodes: In grey are the electrodes selected to obtain the MMN ERP in controls and DS. The electrodes significant between 120-160ms (α ≤ 0.01) were selected following cluster permutation analysis contrasting *standard* and *deviant* trials. The colour bar represent the amplitude of the difference wave obtained subtracting the waveform of *standards* to the waveform of *deviants*.

#### 2.4.4. Cross-sectional statistical analyses

To first assess whether the topography of the ERP MMN differs between the two groups, cluster-based permutations were run for the ERP regardless of condition (merging *standard* and *deviant* trials) and for the dERP. Two-tailed dependent t-tests were used to evaluate the effect between 50ms and 250ms after stimulus onset. At the cluster level the null distribution was generated using the Monte Carlo method and the critical value used for thresholding the sample-specific T-statistics was set at α = 0.01.

The Shapiro-Wilk test for normality was run on all measures and revealed that dERP and dGFP amplitudes and latencies were normally distributed and were retained for all further analyses. However, amplitudes and latencies of ERP and GFP were not normally distributed (all p-values ≤ 0.001) and therefore were omitted from multiple regression analysis.

To assess whether the auditory response to the fifth tone differed between the two groups regardless of condition, ERP and GFP amplitudes and latencies were compared using the non-parametric Wilcoxon rank sum test with continuity correction and to assess whether the MMN differed between the two groups, dERP and dGFP amplitudes and latencies were compared using two-tailed independent samples t-tests. Finally, multiple regressions were fitted to the data and model comparison was conducted to assess whether age predicted changes in MMN across the two groups. Models were built with dERP and dGFP amplitudes and latencies as dependent variables, and with the factors *Group* and *Age* as predictors (models in Formula 1). Models *a* to *d* were compared in the order exposed in Formula 1 using the likelihood ratio test (*lrtest()* in R). Bayes Factors (BF) were also calculated with 50% prior (*bayesfactor_models()* in R). In case of a significant *Group* and *Age* interaction, linear models for the two separate groups were fitted and contrasted to the intercept (models *e* and *f* in Formula 1).

a. *y* ∼1
b. *y* ∼*Age*
c. *y* ∼*Age + Group*
d. *y* ∼*Age ∗ Group*
e. *y*_*group*_ ∼*Age*_*group*_
f. *y*_*group*_ ∼1

**Formula 1** Linear models were fitted and compared to assess whether age predicted changes in MMN across the two groups. *y* is either amplitude or latency of dGFP or dERP. Model a) is the intercept model; model b) expresses the relationship between physiological measures and *Age* as only predictor; model c) includes the main effects of predictors *Age* and *Group*; model d) includes the main effects of the predictors *Age* and *Group* as well as their interaction; model e) expresses the relationship between the dependent variable of only one group (controls or DS) and age; and model f) is the intercept model of data from only one group.

#### 2.4.5. Longitudinal statistical analyses

The analyses focused on the difference between participants’ CAMCOG-DS scores at the cross-sectional phase (Time 1; T1) and the longitudinal phase (Time 2; T2). The total CAMCOG-DS difference score (dCAMCOG) was calculated as the score at Time 2 minus the score at Time 1. To test whether performance at the CAMCOG at T2 decreased compared to the performance at T1, paired t-test was run on CAMCOG scores at the two timepoints. To assess whether changes in CAMCOG-DS scores over time correlate with age, a two-tailed Spearman’s Rank-Order correlation was run between age and dCAMCOG. The Spearman’s test was chosen because research has suggested that the relationship between age and amyloid deposition is not linear (Holland et al., 1998). The BF was calculated for such correlation as well.

Finally we tested whether amplitude and latency of dGFP and dERP at T1 predicted changes in CAMCOG-DS scores over time. We built models with *amplitude* and *latency* of dGFP and dERP as predictors, *IQ* as covariate and dCAMCOG scores as dependent variable. To assess the relationship between variables, linear and curvilinear quadratic models (formula 2) were compared using likelihood ratio tests and BF were calculated.

a. *dCAMCOG* ∼ 1
b. *dCAMCOG* ∼ *x*
c. *dCAMCOG* ∼ *x + IQ*
d. *dCAMCOG* ∼ *x + x*^*2*^ *+ IQ*
e. *dCAMCOG* ∼ *x + x*^*2*^

**Formula 2** Linear (a,b,c) and curvilinear (d,e) models compared to assess whether physiological measures predicted changes in CAMCOG scores (total score and subscales) over time in DS. *x* is either the amplitude or latency of dGFP or dERP.

## 3. Results

### 3.1. Demographics of the cross-sectional phase

Independent samples t-tests for Equality of Means were conducted, with Equality of Variances assumed (p > 0.05) to determine that DS and controls did not significantly differ in age (p = 0.15). Equality of Variance was not assumed (p ≤ 0.05) for group comparisons on hearing acuity, which was determined using the number of tones identified from the Siemens Hear Check Screener. However, an independent samples t-test for the Equality of Means found that the number of tones identified did not significantly differ between groups (p = 0.12). A chi-square test of independence was performed to examine the relationship between gender (male, female) and group (DS, controls) and found no significant relationship: χ^2^(1, 75) = 2.24, p = 0.134. Please see table 1 for more details.

**Table 1.**
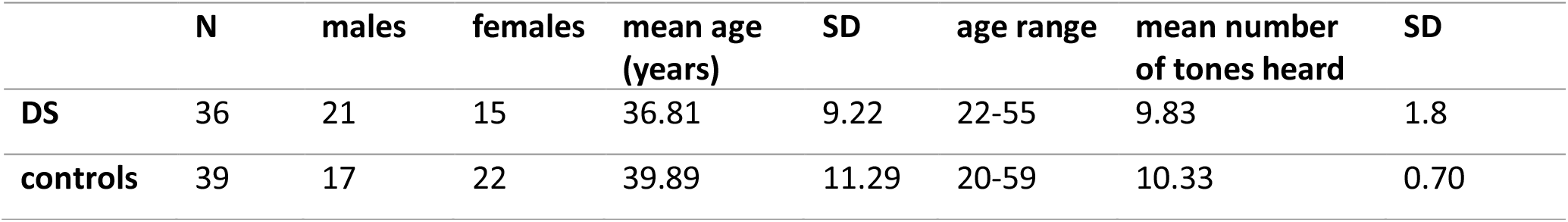
Participant demographics: sex, age and hearing acuity.

|  | N | males | females | mean age<br>(years) | SD | age range | mean number<br>of tones heard | SD |
| --- | --- | --- | --- | --- | --- | --- | --- | --- |
| **DS** | 36 | 21 | 15 | 36.81 | 9.22 | 22-55 | 9.83 | 1.8 |
| **controls** | 39 | 17 | 22 | 39.89 | 11.29 | 20-59 | 10.33 | 0.70 |

Thirty six adults with DS, aged 22-55 years (M = 37.3, SD = 9.39) completed the cross-sectional neuropsychological and EEG testing schedule. Of the 36 adults, 3 had a dementia diagnosis and 21 were male. At the cross-sectional phase, the participants’ total CAMCOG score ranged from 55-105 points (M = 83.1, SD = 13.7). For four participants, KBIT-2 Composite IQ scores could not be used; Verbal IQ scores ranged from 70-80 points (M = 77, SD = 4.7). For the remaining 32 participants, Composite IQ scores range from 40-88 points (M = 53.7, SD = 12.3). The age- and gender-matched controls were aged 20-58 years old (M = 40.08, SD = 11.43), and 17 were male. The lowest Composite IQ score, as assessed by the KBIT-2was 90 (M = 115.76, SD = 22.27); the lowest dementia-screening score, as assessed by the ACE-R, was 88. Therefore, the group is considered to be appropriate as TD controls for this study.

### 3.2. Cross-sectional phase

The cluster permutation run on the ERP regardless of condition between 50ms and 250ms revealed a difference between controls and DS (p ≤ 0.01) at frontal and parietal areas (Fig. 4). The Wilcoxon rank sum test was statistically significant on amplitude (W = 3488, p ≤ 0.01, r = 0.24) and latency (W = 3491.5, p ≤ 0.01, r = 0.24) of the GFP and latency of the ERP (W = 3446, p ≤ 0.01, r = 0.23). DS had smaller amplitude and shorter latency of GFP peaks and shorter latency of ERP peaks than controls (Table 2, Fig. 5). Moreover, when testing for the effect of the MMN in the same time window there was difference between controls and DS (p ≤ 0.01) most pronounced at the right central electrodes (Fig. 4). In DS the dGFP (t_(71.97)_= 4.88, p ≤ 0.001, d = 1.13) and dERP (t_(71.64)_= −2.95, p ≤ 0.01, d = −0.69) peak amplitudes were smaller than in controls. T-tests run on latencies did not show any significant difference in dGFP peaks and only a marginal difference in dERP peaks (t_(71.88)_= −1.94, p = 0.06, d = - 0.45) with DS showing longer latencies (Table 2, Fig. 5).

**Table 2.** Mean and standard deviations (in brackets) of latency (ms) and amplitudes (SD of μV) of ERP and GFP peaks in the two groups regardless of condition and in difference waves.

|  | ERP amplitude | ERP latency | GFP amplitude | GFP latency |
| --- | --- | --- | --- | --- |
| **Controls** | -1.32 (0.92) | 148.74 (11.43) | 1.13 (0.99) | 148.21 (12.04) |
| **DS** | -1.29 (0.97) | 143.69 (11.62) | 0.67 (0.81) | 142.36 (12.92) |

**Table 2** Mean and standard deviations (in brackets) of latency (ms) and amplitudes (SD of $\mu V$ ) of ERP and GFP peaks in the two groups regardless of condition and in difference waves.
|  | dERP amplitude | dERP latency | dGFP amplitude | dGFP latency |
| --- | --- | --- | --- | --- |
| **Controls** | -1.71 (0.67) | 136.74 (9.41) | 1.71 (0.64) | 136.76 (9.41) |
| **DS** | -1.25 (0.68) | 140.78 (8.54) | 1.00 (0.60) | 137.83 (10.04) |

**Figure 4.**
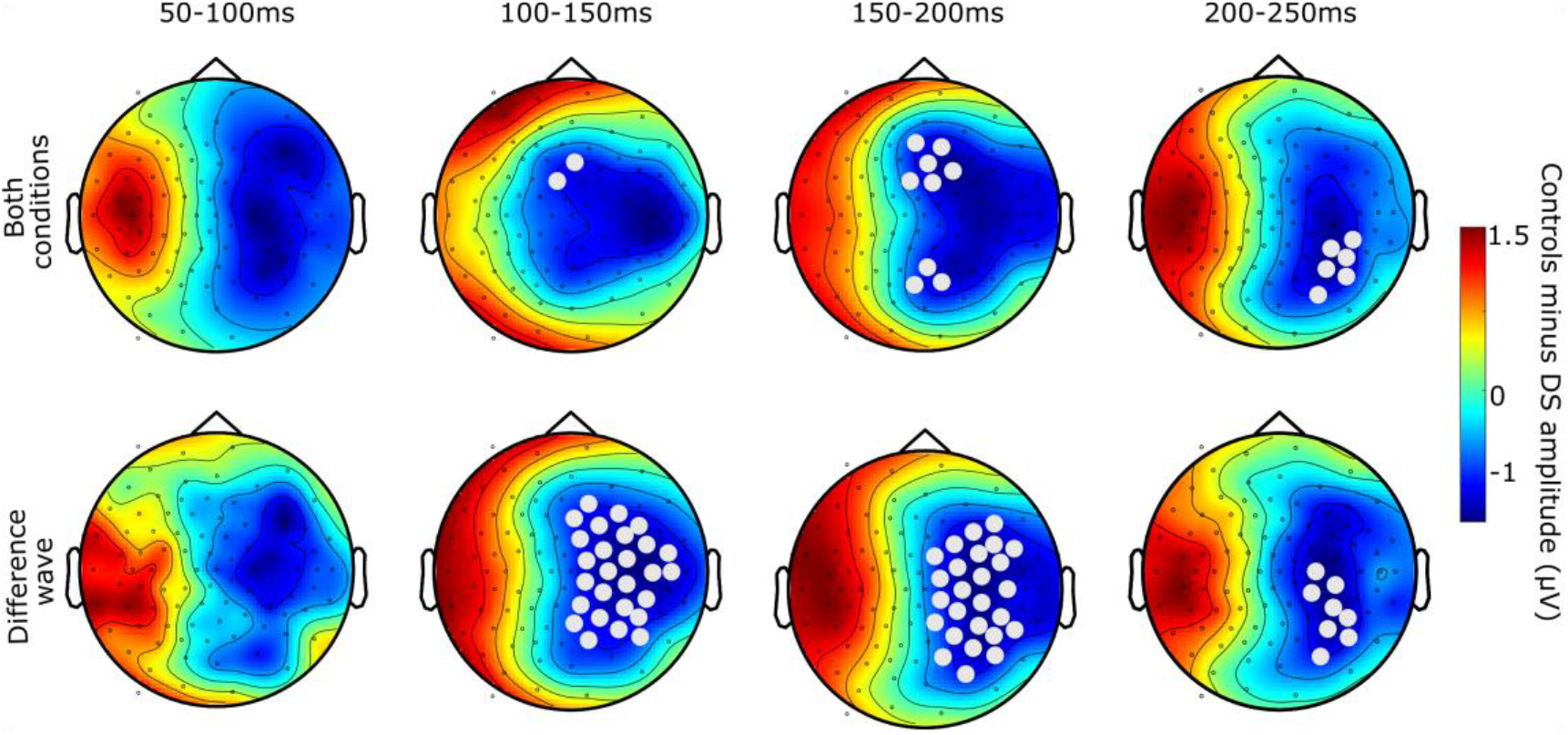
Cross-sectional cluster permutation: Results of cluster permutation analysis between 50-250ms. In the top row are plotted the results of the analysis of the ERP of *standard* and *deviant* conditions combined contrasting DS and controls. In the bottom row are plotted the results of the analysis of the *difference waves* DS and controls. Significant electrodes (α ≤ 0.01) are marked in grey. The colour bar represent the amplitude of the waveform obtained subtracting the waveform of DS to the waveform of controls.

**Figure 5.**
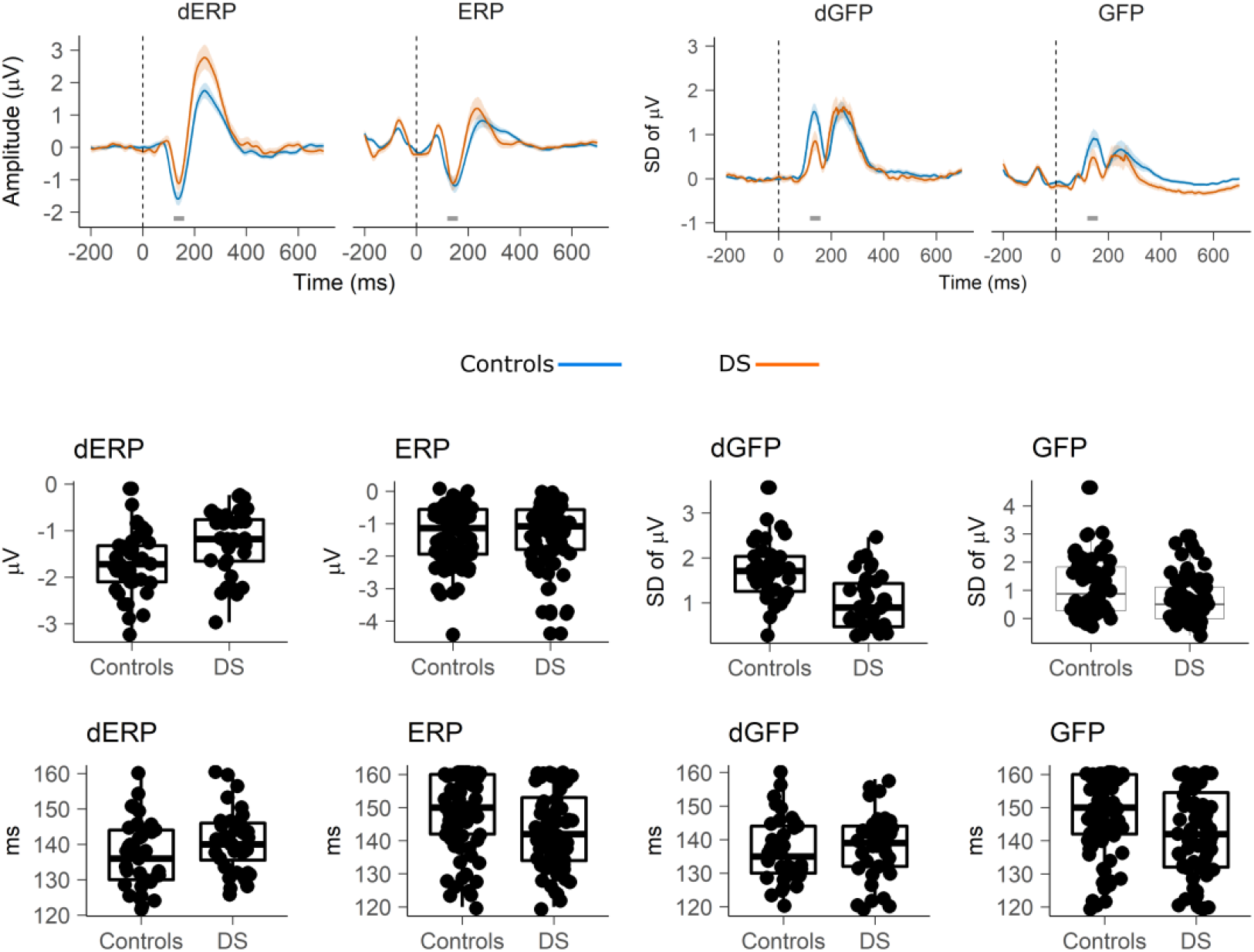
Cross-sectional results: Waveforms (top row), boxplots representing peak amplitudes (second row) and peak latencies (bottom row) of dERP (first column), ERP regardless of condition (second column), dGFP (third column), and GFP regardless of condition (fourth column) of controls and DS. In the waveform plots, the grey segment represents the 120-160ms window in which the peaks have been identified, shading represents 95% confidence intervals.

Comparing the models that predicted dGFP amplitudes based on *Age* and *Group*, the model with *Group* and *Age* (model *c* in Formula 1) provided a significant improvement compared to the model (model *b* in Formula 1) with *Age* as only predictor (χ^2^_(1)_ = 20.78, p ≤ 0.001, λ = −68.84, BF_cb_ = 3780). The model with the interaction between *Age* and *Group* (model *d* in Formula 1) was significantly better (χ^2^_(1)_ = 7.54, p ≤ 0.01, λ = −65.07, BF_db_ = 5.05) than the model with only the main factors (model *c*). Model *d* explained a significant amount of the variance of dGFP amplitude changes with age (F_(3,70)_ = 11.02, p ≤ 0.001, R^2^ = 0.32, R^2^_Adj_ = 0.29). Of the coefficients, only the interaction statistically predicted dGFP amplitude (B = −0.04, SE = 0.01, t_(70)_ = −2.74, p ≤ 0.01). Confirming that age alone was not predictive of dGFP amplitudes, the BF_ba_ = 0.14 between model b and the intercept (model *a*) was obtained. The linear model fitted only on TD data (model *e*, Formula 1) was not significant and did not differ from the intercept (model *f*, Formula 1) as confirmed by BF_ef_ = 0.54, suggesting that *Age* is not a good predictor of dGFP amplitude in TD. On the other hand, the model fitted on DS data (model *e*) was significant (F_(1,34)_ = 5.57, p ≤ 0.05, R^2^ = 0.14, R^2^_Adj_ = 0.12). The predictor *Age* significantly predicted dGFP amplitude in DS with smaller amplitudes in older individuals (B = −0.02, SE = 0.01, t_(35)_ = −2.36, p ≤ 0.05) and the model differed from the intercept (χ^2^_(1)_ = 5.46, p ≤ 0.05, λ= −29.28, BF_ef_ = 2.56). The loglikelihood test on dGFP latencies found no improvements of the alternative models compared to the intercept. When running incremental bayesian model comparisons, the simpler model was always probable with a greater degree of belief than the alternative, more complex model (BF_ba_ = 0.21, BF_cb_ = 0.14, BF_db_ = 0.12).

Regarding dERP, comparing the models that predicted variations in amplitudes based on *Age* and *Group*, the model with *Group* and *Age* (model *c* in Formula 1) provided a significant improvement to the model (model *b* in Formula 1) with *Age* as only predictor (χ^2^_(1)_ = 8.45, p ≤ 0.01, λ= −75.00, BF_cb_ = 7.96). The model with the interaction between *Age* ad *Group* (model *d* in Formula 1) was significantly better (χ^2^_(1)_ = 11.01, p ≤ 0.001, λ= −69.50, BF_db_ = 28.55) than the model with only the main factors (model *c*). Model *d* explained a significant amount of the variance of dERP amplitude changes with age (F_(3,70)_ = 7.04, p ≤ 0.001, R^2^ = 0.23, R^2^_Adj_ = 0.20). In the model, the predictor *Group* (B = −1.42, SE = 0.58, t_(70)_ = −2.43, p ≤ 0.05) and the *Group* and *Age* interaction (B = 0.05, SE = 0.01, t_(70)_ = 3.35, p ≤ 0.01) significantly predicted dERP amplitude. The predictor *Age* was not significant but approached significance (B = −0.02, SE = 0.01, t_(70)_ = −1.91, p = 0.06). To further investigate the role of age regardless of group, the BF_ba_ = 0.12 between model *b* and the intercept (model *a*) was obtained. The linear model fitted only on control data (model *e*, Formula 1) was not significant and did not differ from the intercept (model *f*, Formula 1) as confirmed by BF_ef_ = 0.96. On the other hand, the model fitted on DS data (model *e*) was significant (F_(1,34)_ = 7.94, p ≤ 0.01, R^2^ = 0.19, R^2^_Adj_ = 0.17). The predictor *Age* significantly predicted dERP amplitude in DS (B = 0.03, SE = 0.01, t_(35)_ = 2.82, p ≤ 0.01) and the model differed from the intercept (χ^2^_(1)_ = 7.56, p ≤ 0.01, λ= −33.00, BF_ef_ = 7.31). The loglikelihood test on dERP latency showed that model *c* with the main factors had better fit than model *b* (χ^2^_(1)_ = 5.27, p ≤ 0.05, λ = −264.10, BF_cb_ = 1.62). Model *c* explained a significant amount of the variance of dERP latency (F(2,71) = 4.41, p ≤ 0.05, R^2^ = 0.11, R^2^_Adj_ = 0.09) and both *Age* (B = 0.22, SE = 0.10, t_(71)_ = 2.21, p ≤ 0.05) and *Group* (B = 4.72, SE = 2.06, t_(71)_ = 2.29, p ≤ 0.05) were significant predictors of latency. Hence, the data showed that latency slowed down as a function of age in both groups. To further explore the role of age, model b was compared against the intercept and a BF_ba_ = 0.64 showed only anecdotal evidence for model *b*. Multiple regressions are plotted in Fig. 6.

**Figure 6.**
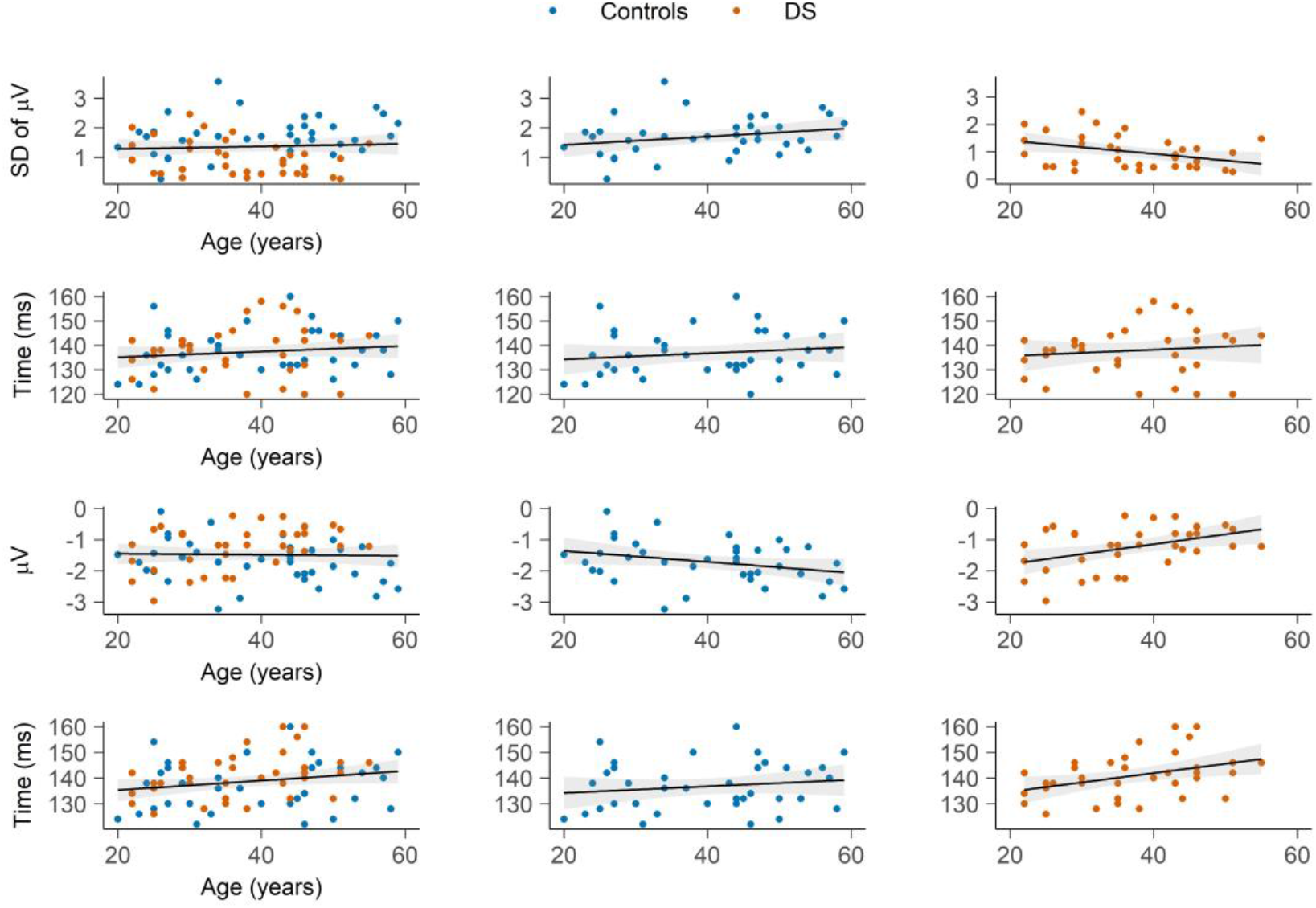
Prediction of age: Fitted linear models with dGFP amplitude (first row), dGFP latency (second row), dERP amplitude (third row), and dERP latency (fourth row) as dependent variable and *age* as predictor. In the second and third column, the linear models have been plotted separately for the *group* factor so that to illustrate the interaction between *age* and *group* predictors. Shading represents 95% confidence intervals.

### 3.3. Demographics of the longitudinal phase

Thirty-four of the 36 adults with DS completed both the initial (cognitive and EEG) and the follow-up (cognitive only) assessments. One participant was deceased and one participant was excluded because had fulfilled criteria for “uncooperative behavior”, “silly behavior” and “flat affect”. Of the 34 adults, 3 had a dementia diagnosis at time 1 (T1), 20 were male and 32 were right handed. No participants transitioned to an AD diagnosis between T1 and T2. Table 3 provides more demographic and cognitive detail for the cognitive follow-up of participants.

**Table 3.** Demographics of the longitudinal phase participants. T1 represents time 1 (initial assessment), T2 represents time 2 (follow-up assessment). SD is standard deviation from the mean.

|  | min | max | mean | SD |
| --- | --- | --- | --- | --- |
| **T1 age (years)** | 22 | 55 | 37.6 | 9.4 |
| **T1 total CAMCOG score** | 58 | 105 | 83.4 | 14.0 |
| **T2 total CAMCOG score** | 48 | 104 | 81.4 | 15.1 |
| **T2-T1 total CAMCOG score** | -17 | 5 | -2.0 | 4.7 |

### 3.4. Longitudinal phase

Scores on the CAMCOG at T2 were significantly lower than scores at T1 (t_(34)_ = 2.51, p≤0.05, Cohen’s d = 0.42). The Spearman’s Rank-Order correlation between dCAMCOG and age was not significant (ρ(33) = −0.16, p = 0.36, BF_10_ = 1.07. Fig. 7). Therefore, age was considered no further in the following analyses. The 34 participants’ total CAMCOG difference scores ranged from −17 to 5 points with a mean change of −2.06 points (SD = 4.77 points). Sequentially comparing the models reported in Formula 2 with dGFP amplitude as predictor, no model was a significant improvement over the preceding model (BF_ba_ = 0.31, BF_cb_ = 0.17, BF_dc_ = 0.21, BF_de_ = 0.17), suggesting that dGFP amplitude was not a predictor of cognitive decline as measured by dCAMCOG scores. The same model comparison on models with dGFP latency as predictor, showed that the curvilinear model d was significantly better than the linear model c (Loglikelihood = −98.67, χ^2^(1) = 5.83, p ≤ 0.05, BF_dc_ = 3.11). However, model *d* did not have a significantly better fit than model *e*. Therefore model *e* was kept as the chosen model being a simpler model. Model *e* was statistically significant (F_(2,32)_ = 4.79, p ≤ 0.05, R^2^ = 0.23, R^2^_Adj_ = 0.18) and both the linear (B = 3.87, SE = 1.60, t_(32)_ = 2.42, p ≤ 0.05) and quadratic (B = −0.01, SE = 0.006, t_(32)_ = −2.33, p ≤ 0.05) predictors were significant in the model (Fig. 7), which suggested that there is a nonlinear relationship between dGFP latencies and cognitive decline as measured by dCAMCOG scores. Specifically, bigger cognitive decline (smaller dCAMCOG scores) predicted shorter and longer latencies while intermediated dCAMCOG scores predicted intermediate latencies. No model was a significant improvement over the preceding model in models with dERP amplitude as predictor (BF_ba_ = 0.21, BF_cb_ = 0.17, BF_dc_ = 0.17, BF_de_ = 0.17) and in models with dERP latency as predictor (BF_ba_ = 0.17, BF_cb_ = 0.17, BF_dc_ = 0.18, BF_de_ = 0.17).

**Figure 7.**
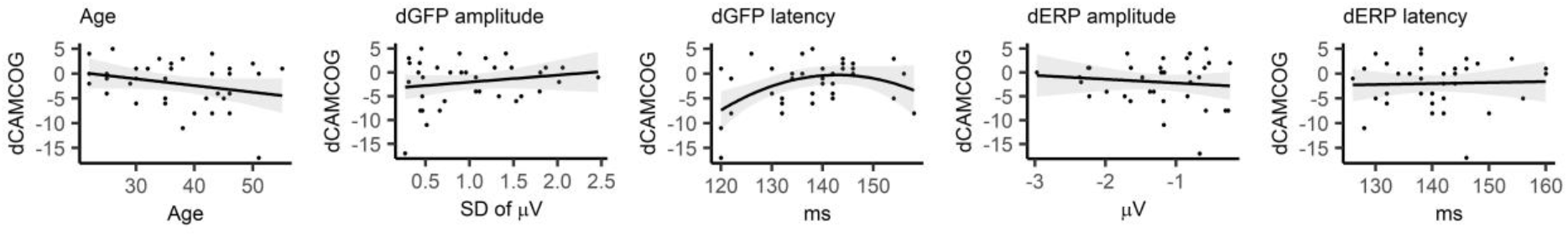
Prediction of cognitive decline: Correlation between total dCAMCOG score and age (first plot). Linear models with dGFP amplitude, dGFP latency, dERP amplitude, and dERP latency as predictors and total dCAMCOG score as dependent variable (second to fifth plot). Shading represents 95% confidence intervals.

The curvilinear quadratic model was then fit to dGFP latency data parsed in CAMCOG subscales where the T2-T1 score to each subscale was set as dependent variable. The BF against the intercept was also obtained. The model was not significant for any of the subscales (Table 4).

**Table 4.** T2-T1 difference scores to the CAMCOG subscales. Means (M) and standard deviations (SD) are reported. BFs are calculated comparing the curvilinear model e (Formula 2) against the intercept (model a in Formula 2). BF are expressed as the alternative model (1) against the denominator model (0).

| Subscale | M | SD | BF<sub>10</sub> |
| --- | --- | --- | --- |
| orientation | -0.03 | 1.29 | 0.05 |
| language | 0.06 | 2.71 | 0.04 |
| memory | 0.11 | 2.91 | 0.08 |
| attention | -0.23 | 1.11 | 0.05 |
| praxis | -0.57 | 1.61 | 0.34 |
| perception | 0.43 | 1.15 | 0.03 |
| abstract thinking | -1.17 | 2.38 | 0.07 |

## 4. Discussion

Individuals with DS experience premature ageing and AD pathology affects nearly the entirety of the DS population with increasing age (Grothe et al., 2017; Sperling et al., 2013; Teipel et al., 2020). The prolonged preclinical phase of AD during which time neuropathology is developing indicates the importance (Näätänen et al., 2004) of identifying early markers of cognitive decline to both enable the assessment of potential treatments and also to indicate in whom and when treatments once developed should be started. The present research aims to identify electrophysiological markers of premature ageing in DS and predictors of cognitive decline.

To assess premature ageing in DS, DS and TD age-matched controls were presented sequences of five tones that could either consist of five identical tones (*standard*) or four identical tones and a fifth different tone (*deviant*). The contrast between standard and deviant tones generate the MMN component, which is believed to be a marker for the brain’s automatic detection of auditory regularity violation (Näätänen et al., 2011). The MMN depends on temporo-frontal short-term memory trace in the auditory cortex (Cheng et al., 2013). Hence, the decline in sensory memory, perceptual accuracy and the brains’ predictive power with ageing is reflected in decreased MMN response (Näätänen et al., 2011). To assess whether the MMN is a predictor of cognitive decline in DS, CAMCOG scores were obtained at the same time as the EEG recording (T1) and 10 to 14 months later (T2) for DS participants. The relationship between the MMN at T1 and the CAMCOG difference score between the two timepoints allowed on the predictive power of the MMN to be studied. Both traditional ERP and whole scalp GFP techniques were employed to address these research questions. The MMN is considered an index of cognitive decline in several neuropsychological disorders (Lalo et al., 2005). However, it has not been extensively studied in participants with DS (Romano et al., 2016). Hence, we first compared DS and controls in the response elicited by the fifth tone regardless of condition and in the difference wave (*standard* minus *deviant*) independently of age. dERP and dGFP, which directly represents the MMN marker of auditory regularity violation, consistently showed a reduced MMN response in DS compared to controls (Fig. 5). It is possible that such difference is driven by age-related reduction in grey matter which is apparent already at 20 years of age (Näätänen et al., 2011) and that may impair the ability of forming sensory memory traces. The difference in MMN responses between groups is widespread over central-right electrodes (Fig. 4). Because interaural stimulation was counterbalanced between left and right ear in both groups, the topography is not an artifact of stimulus presentation. In fact, it may indicate that DS are mostly impaired in the involuntary attention switch to auditory change, which has been found to be a right-lateralised process (Näätänen & Picton, 1987; Tomé et al., 2015).

When looking at the response to the fifth tone regardless of condition, the cluster permutation revealed that the ERP response of DS and controls differed over localized frontal and parietal electrodes. It is possible that such effect is driven by the response to *deviant* sequences. However, it is also possible that the cluster permutation detected differences in the N1 component that overlaps with the MMN (Tomé et al., 2015). The N1 is a bilateral frontocentral component elicited by both frequent and infrequent stimuli and reflects sensory and perceptual processes (Näätänen & Picton, 1987; Tomé et al., 2015) suggesting that DS may present sensory and perceptual impairment regardless of abstract detection of regularity violation. We also compared DS and controls on the ERP peaks that were extracted only from the electrodes that had formed significant clusters when contrasting *standard* and *deviant* sequences (Fig. 3). Peak comparison yielded no differences in amplitude between DS and controls. It has been suggested that several different components generated by different cerebral locations and sub-serving different cognitive processes contribute to a widespread negativity peaking around 150ms (Tomé et al., 2015). The electrodes selected may have recorded the summation of such processes and did not measure any difference in response intensity to tones between the two groups. We found key differences in GFP amplitude, where DS had smaller N1 responses compared to controls. GFP is a whole-scalp measure, so the GFP might have picked up the group difference at frontal electrodes underlying the same frontal N1 effect emerged in the cluster permutation analysis. Surprisingly, latencies were reduced in DS compared to controls in both GFP and ERP. Findings on the N1 latency have been inconsistent (Eggermont, 1988; Ponton et al., 2000; Tomé et al., 2015), but several studies, which have reported small latency decreases with advancing age, have attributed shorter N1 to increased synaptic synchronization and efficiency during brain maturation (Horváth et al., 2007; Kiang et al., 2009; Schiff et al., 2008). This seems an unlikely scenario in this case.

The models predicting dGFP and dERP amplitude as a function of age and group show that amplitude decreases with age in DS but not in controls. In a predictive coding framework, the results suggest that the ability of making accurate statistical predictions about the incoming stimuli decreases with age in DS but not in controls. In DS, the neurodegeneration caused by Aβ depositions over time may compromise anatomical connections between the nodes of the frontotemporal network, reducing the connectivity between these areas and compromising accurate prediction. According to models of the generation of the MMN (Friston, 2003, 2005; Garrido et al., 2008; Garrido et al., 2009a; 2009b), the prediction error represented by the MMN is dependent on both backward (top-down) and forward (bottom-up) connections between the levels of the network hierarchy. Future studies should aim at identifying how age-related decay of the MMN response in DS affects these two directions of connectivity.

On the other hand, amplitudes seem not to be affected by the main effect of age. Notably, while the main factor *Age* was significant when it was the only predictor of dERP amplitude, the BF of 0.64 shows that the model cannot be taken as conclusive evidence of age-related MMN decay. The effect of age on MMN amplitude had been hypothesized for both groups based on the evidence that attenuated amplitude and longer latencies have been systematically found in normal ageing (Cheng et al., 2013; Näätänen et al., 2011, 2014). However, in our studies DS and controls had an average age of 35 and 40 years respectively, while studies assessing the MMN in elderly controls typically have older participants. It may be that the sensory memory decay in our cohort was not sufficient to be reflected in changes in the MMN. This is consistent with the hypothesis that at 40 years of age, DS begin to show abnormal Aβ binding in the brain compared to controls (Annus et al., 2016). Interestingly, the absence of an effect of age in typically developing controls suggests that variation in the MMN may be a sensitive measure of accelerated ageing that is able to distinguish between pathological and normal ageing.

In contrast to the results on amplitude, there is no effect on MMN latency. This is not surprising as evidence for the effect of ageing on MMN latency have been inconsistent (Annus et al., 2016; Holland et al., 1998; Lautarescu et al., 2017). One interpretation might be that sensory memory in DS is reduced with age but not necessarily slowed down, sensory information may simply not be properly encoded. This hypothesis needs further assessment. Overall, these results support the hypothesis that the amplitude of the MMN is a potential marker of accelerated ageing in DS. The cohort of DS participants included three individuals with AD. The decision to keep those participants in our sample was driven by the strong evidence that DS will invariantly develop AD during their lifespan (Näätänen et al., 2011). The presence of AD is typically considered a dichotomous variable used as inclusion or exclusion criterium. Instead, we made the theoretical decision of treating it as a continuous variable that simply expresses a degree of progression of inevitable pathological ageing in individuals with DS.

Cross-sectional data suggest that the MMN manifested in both GFP and ERP measures is a measure sensitive to DS accelerated ageing. However, its relationship with cognitive decline over time is less clear. First, there was no correlation between dCAMCOG and age. Second, there only seem to be moderate evidence for a nonlinear relationship between dGFP latency and dCAMCOG scores, with increasing latency predicting stronger cognitive decline (lower dCAMCOG scores) at shorter and longer latencies but weaker decline at intermediated latencies. Subsequently, no relationship was found between dGFP latency and CAMCOG subscales difference scores. Given the results, there is little evidence for any relationship between MMN measures and cognitive decline as measured through the CAMCOG. The contrast between scores at T1 and T2 showed that the CAMCOG was able to detect a significant cognitive decline after 12 months, although with a small effect size. Hence, one explanation of a lack of relationship between MMN and dCAMCOG may be that the extent of cognitive decline was not big enough to be reflected in MMN variations. One reason for this may be that the time gap between T1 and T2 might not have been long enough to see a statistically significant effect. Alternatively, the study might be underpowered to detect potentially small effects. It is also important to keep in mind that the CAMCOG assesses a variety of complex cognitive functions. On the other hand, smaller amplitudes of the pre-attentive MMN reflect decline in sensory memory (Horst et al., 1993; Huppert et al., 1996) which is a lower level function. Hence, the complexity of the CAMCOG scores may overshadow any decline on sensory memory. Finally, one may argue that if sensory memory declines then we could have expected the MMN to predict scores on the Perception subscale. However, perception is measured through recognition of famous people and objects from different angles (Horst et al., 1993; Huppert et al., 1996) and thus sensory memory is not directly assessed.

A limitation of the present research is that there was no MMN measurement at T2. Future research should aim at filling this gap. First, it should assess the degree of MMN variation over time in DS within individual participants. If the MMN is a potential tool to measure accelerated ageing in the DS population, it is clinically important to establish its temporal sensitivity. Furthermore, the MMN at T1 may not be predictive of cognitive decline after 12 months but a change in MMN amplitude over time may be predictive of future cognitive decline at a larger timescale. Finally, the possibility of MMN latency variations as a measure of accelerated ageing should be further investigated.

## 5. Conclusions

The present study investigated the MMN as a means to study accelerated ageing in DS. We showed that age predicted the amplitude of the MMN both expressed in GFP and traditional ERP. The amplitude decreased with increasing age. On the other hand, in the age-matched typically developing group, age did not predict MMN amplitude. This suggests that people with DS electrophysiologically show accelerated ageing and that the MMN is a valid marker to assess the impairment of cortical processes of sensory memory and structures associated with the generation of MMN responses.

The study also investigated the MMN as predictor of cognitive decline at about 12 month time. Overall the data did not provide strong evidence in support of a significant relationship between MMN and cognitive decline in DS as measured by the CAMCOG-DS - neither for the total score, or for its subscales. Future research should look at the MMN longitudinally and should investigate whether rates of MMN change over time predict cognitive decline.

## Data Availability

The data that support the findings of this study are available from the corresponding author, CA, upon reasonable request.

## Acknowledgements

We are very grateful to the adults who participated in the study, and their families and carers who supported them to do so. The Medical Research Council, The Health Foundation, Addenbrooke’s Charitable Trust, Alzheimer’s Research UK Pump Priming Grant – Cambridge Network, Fearnsides Fund, and Marmaduke Sheild Fund funded this research. We thank them for their support. We appreciate the assistance from the DSA and JDR in identifying participants for the study. We also would like to thank Paula Castro for her contribution with data collection and Dr Peter Watson for valuable statistical advice.

## Authors contribution

Conceptualization and design: SJ, TB, VN, SC, AH, SZ, HR. Data acquisition: SJ, MW, PC. Data analysis: CA, SJ. Drafting: CA, SJ. Figures: CA. Supervision: SC, VN, TB, HR. Review: CA, AL, VN, AH, IC. Funding: AH, SZ, HR, SJ. CA and SJ contributed equally to the paper.

